# Social Distancing Causally Impacts the Spread of SARS-CoV-2: A U.S. Nationwide Event Study

**DOI:** 10.1101/2020.06.29.20143131

**Authors:** Louis Gagnon, Stephanie Gagnon, Jessica Lloyd

**Author notes:** Corresponding author. Louis Gagnon is a Professor of Finance at the Smith School of Business, Queen’s University, Kingston, Ontario, Canada, K7L 3N6.

## Abstract

**Objectives:** To assess the causal impact of a spontaneous relaxation of social distancing on the spread of SARS-CoV-2 in the United States (U.S.), while controlling for social mobility and state-imposed social distancing restrictions.

**Design:** Event study.

**Setting:** Quasi-experimental setting created by the U.S. nationwide protests precipitated by George Floyd’s tragic death on May 25, 2020.

**Population:** Individuals in 3,142 U.S. counties from all 50 states and the District of Columbia.

**Main Outcome Measures:** The number of daily confirmed COVID-19 cases in all U.S. counties between the period of January 22, 2020, and June 20, 2020, and the cumulative change in COVID-19 cases in protest counties relative to non-protest counties following the onset of the protests.

**Results:** We document a country-wide increase of over 3·06 cases per day, per 100,000 population, following the onset of the protests (95%CI: 2·47–3·65), and a further increase of 1·73 cases per day, per 100,000 population, in the counties in which the protests took place (95%CI: 0·59–2·87). Relative to the week preceding the onset of the protests, this represents a 61·2% country-wide increase in COVID-19 cases, and a further 34·6% increase in the protest counties.

**Conclusions:** Our study documents a significant increase in COVID-19 case counts in counties that experienced a protest, and we conclude that social distancing practices causally impact the spread of SARS-CoV-2. The observed effect cannot be explained by changes in social distancing restrictions and social mobility, and placebo tests rule out the possibility that this finding is attributable to chance. Our research informs policy makers and provides insights regarding the usefulness of social distancing as an intervention to minimize the spread of SARS-CoV-2.

## 1 Introduction

The highly contagious novel coronavirus, severe acute respiratory syndrome coronavirus-2 (SARS-CoV-2), responsible for coronavirus disease 2019 (COVID-19), emerged in December 2019 in Wuhan city, Hubei province, China. ^1^ The initial COVID-19 outbreak quickly evolved into a pandemic, ^2^ and as of June 2020, SARS-CoV-2 has reached over 180 countries and regions, with the total number of confirmed cases surpassing 10 million globally.^3^ COVID-19 has spread throughout the United States (U.S.) at an unparalleled rate, infecting over 2·5 million individuals and claiming over 125,000 lives.^4^ Global public health measures aimed at reducing the spread of SARS-CoV-2 have been designed in consideration of the virus’s specific transmission properties. ^5^ SARS-CoV-2 can be transmitted through various modes, including person-to-person contact and the spread of respiratory droplets, which can travel across a minimum distance of 6 feet (2 meters). ^6,7^ Numerous countries have introduced social distancing, defined as the maintenance of at least a 6 foot interpersonal physical separation, to minimize direct transmission from infected individuals. ^8^

In the U.S., individual states have been granted the authority to design their own COVID-mitigation strategy, therefore, the extent and type of social distancing policies adopted differs across states. ^9^ Research examining state-imposed restrictions has found a reduction in the doubling rate of SARS-CoV-2 among U.S. states,^10^ as well as the daily growth rate of COVID-19 cases across U.S counties after the imposition of social distancing measures. ^11,12^ Other research has suggested that rather than reducing the daily growth rate of COVID-19, social distancing merely stabilizes the spread of SARS-CoV-2 in the U.S. ^10^ Additionally, when examining the effectiveness of social distancing, studies have used social mobility as a measure of social distancing.^11,13–15^ However, mobility represents an imperfect proxy for social distancing because individuals can be mobile while still maintaining the required minimum 6 foot separation from others to prevent viral transmission. Furthermore, although evidence suggests there is an association between social distancing and the spread of SARS-CoV-2, the causal impact of social distancing on the spread of SARS-Cov-2 is still unknown.

In this study, we examine the the causal impact of a spontaneous relaxation of social distancing measures on the spread of SARS-CoV-2. The nationwide mass protests precipitated by George Floyd’s tragic death on May 25, 2020 prompted an abrupt relaxation of social distancing practices across the U.S. The unpredictable nature of the protests created a natural experimental setting to assess for causality. In this study, instead of using mobility as a proxy for social distancing, we control for the increase in mobility during the protest period in order to hone in on the direct effect of social distancing. We also explicitly control for the concurrent relaxation of state-imposed restrictions to account for variations in social distancing restrictions across states.

## 2 Methods

### 2.1 Data and Sample Description

We source our U.S. COVID-19 data from the John Hopkins GitHub repository. This data consists of confirmed cases in each county at the end of every day since the start of the outbreak in late January 2020. We calculate the number of new cases for each county and each day by subtracting the cumulative number of confirmed cases at the end of the day from the number of cumulative cases from the previous day. This sampling procedure yields a panel data-set consisting of a total of 474,422 county-days representing 3,142 counties from all fifty states, as well as the District of Columbia (DC), for the period starting on January 22, 2020, and ending on June 20, 2020. We describe our sample in Table I, and in Figure 1 we show the counties in which protests took place according to media reports, along with the size of the first protest taking place within each county.

We obtain our county-level population data and our county-level demographic data from the U.S. Census Bureau. We extract our county-level Gross Domestic Product (GDP) data from the U.S. Bureau of Economic Analysis’ (BEA) Regional Economic Accounts database (Table CAGDP1). We retrieve county-level data on the prevalence of obesity, diabetes, smoking, and hypertension from the University of Washington’s Institute for Health Metrics and Evaluation (IHME). The hypertension and obesity data are for the years 2009 and 2011, respectively, and the diabetes and smoking prevalence data are for 2012. The IHME reports hypertension and obesity data for females and males separately, so we construct a population-weighted average measure for these two covariates based on the proportion of females and males in each county, as reported by the U.S. Census Bureau.

The social distancing restrictions data is from the University of Washington’s State-Level Social Distancing Policies in Response to the 2019 Novel Coronavirus in the U.S. repository. The social distancing restrictions include: 1) restrictions on public gatherings exceeding 5, 10, 25, 50, 100, 250, 500, or 1,000 people, 2) limits on restaurant operations, 3) closure of specific businesses, e.g. fitness centres, gyms, casinos, etc., 4) closure of non-essential businesses, 5) stay-at-home orders for non-essential activities, 6) state curfews on non-essential activities, 7) mandated quarantines for people entering the state, 8) travel restrictions prohibiting residents from leaving the state, non-residents from entering the state, or residents from travelling across counties within the state, 9) self-isolation requirement for individuals with confirmed COVID-19 infection, and 10) mandatory wearing of masks or other mouth and nose coverings in public places. We construct our social distancing restrictions index by adding the number of restrictions that are in place in a state on any given day, based on the date at which each restriction is enacted, relaxed, or expired. Figure 2 shows the evolution of our index for randomly selected states.

**Table I:**
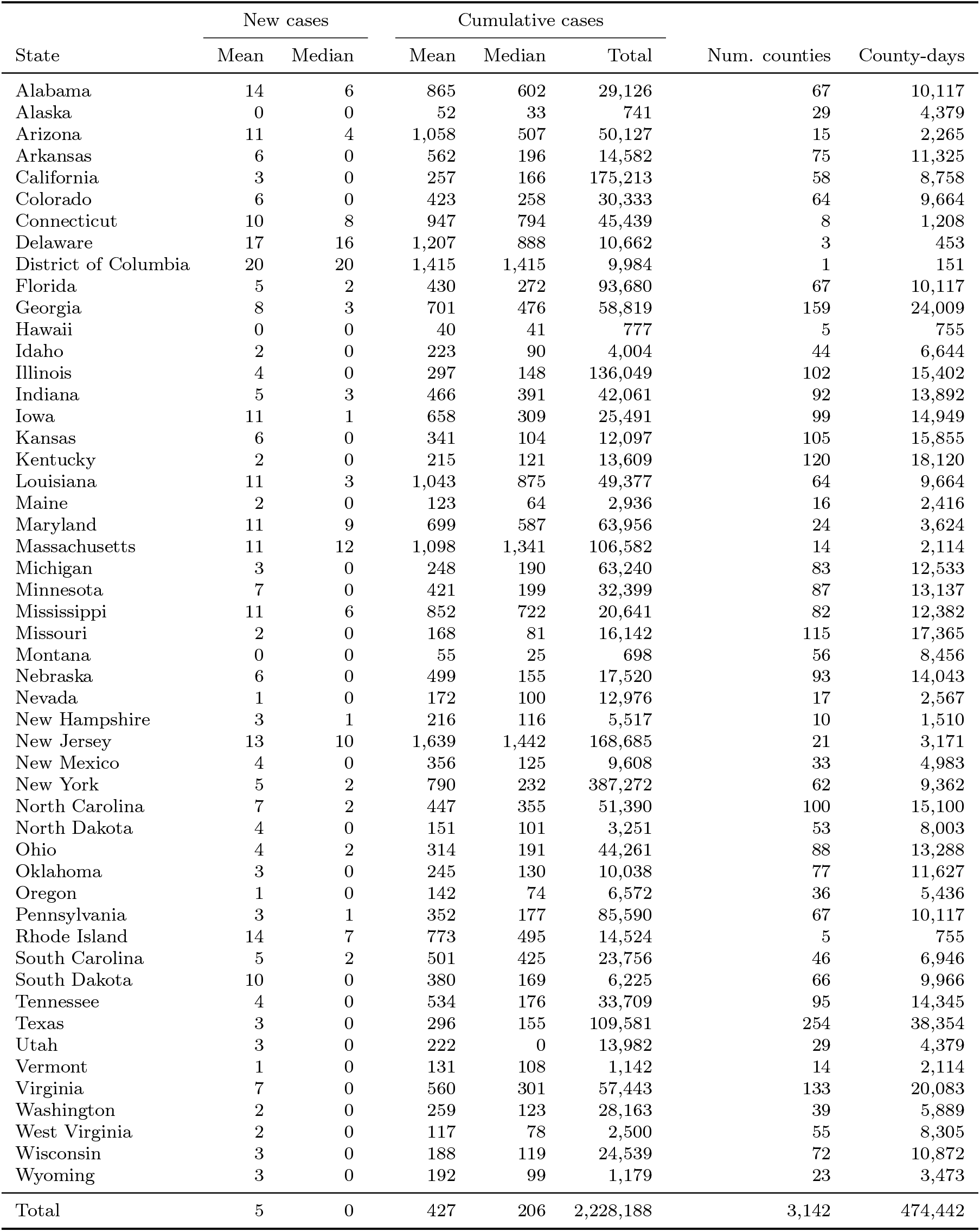
Sample Description. This table reports the mean and the median number of new COVID-19 cases, per day, per 100,000 population, during the week preceding the onset of the protests (May 18-24, 2020), as well as the mean, median, and total number of confirmed cases, across all counties within each state at the end of our sample period. The number of counties and county-days represented in our sample within each state are reported in the last two columns of the table. Our sample period begins on January 22, 2020, and ends on June 20, 2020.

**Figure 1:**
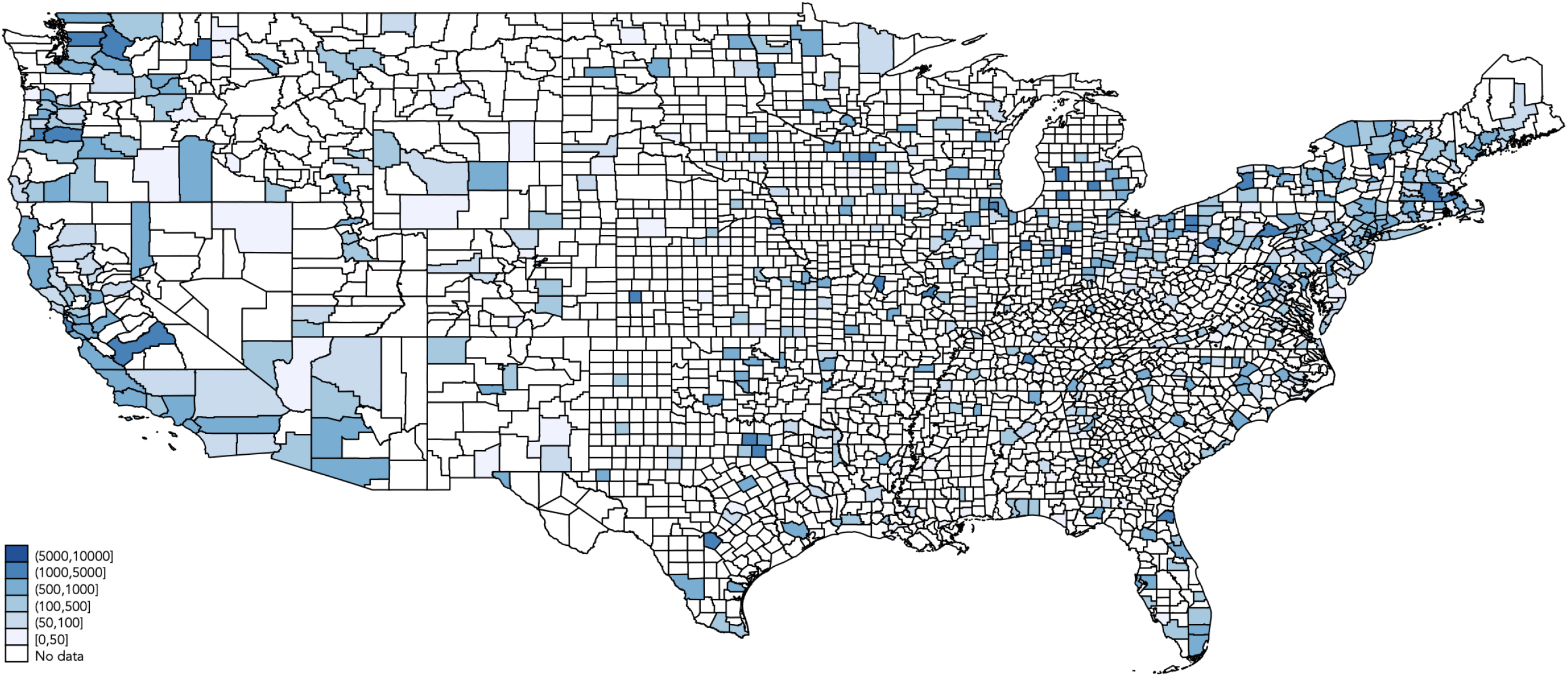
Counties Involved in Protests. This figure identifies the counties in which protests took place, according to media reports, along with the number of participants involved in the first protest that took place within each county. Counties within the states of Alaska and Hawaii are not shown, but they are included in our sample.

**Figure 2:**
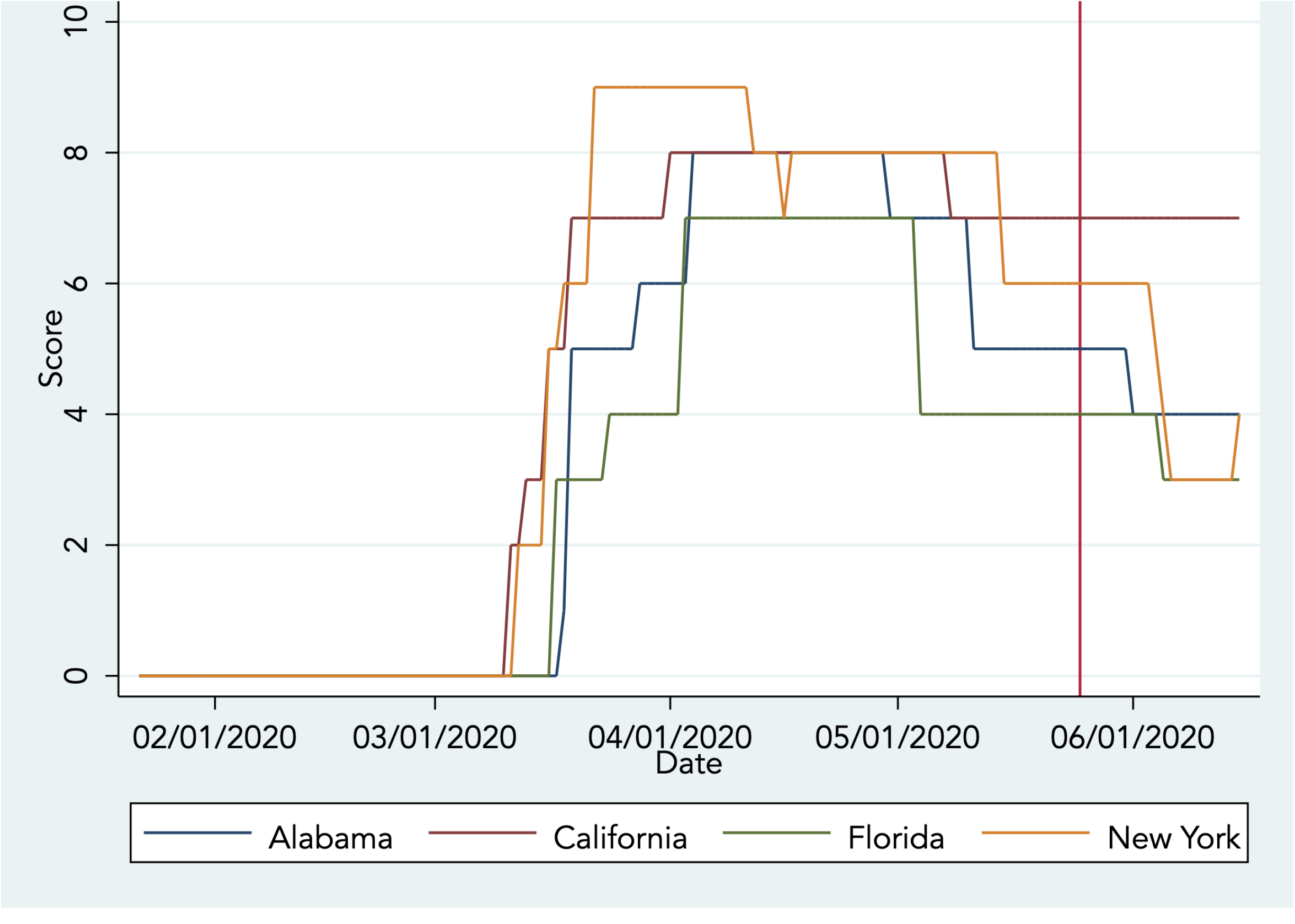
Social Distancing Restrictions Index. This figure shows the evolution of our social distancing restrictions index during our sample period for the states of Alabama, California, Florida, and New York. We selected these states randomly for illustrative purposes only. The vertical line corresponds to May 26, 2020, the day of the onset of the protests.

We obtain our mobility data from the Descartes Labs. This data consists of mobility indexes calculated at the end of every day and aggregated at the county level. The indexes, which we will refer to as the social mobility index, are based on geolocation reports from smartphones and other mobile devices, and track the movements of individual mobile phone subscribers. The methodology employed to construct these indexes is described in Warren et al., 2020. ^16^ The mobility index data is available at a daily frequency from March 1, 2020, until the end of our sample period. Thus, we lose a total of 122,538 county-day observations from the start of our sample period up until February 29, 2020, in all our regression analyses featuring this data. Figure 3 shows the mobility index for a randomly selected small and large county in the states of New York and Texas.

**Figure 3:**
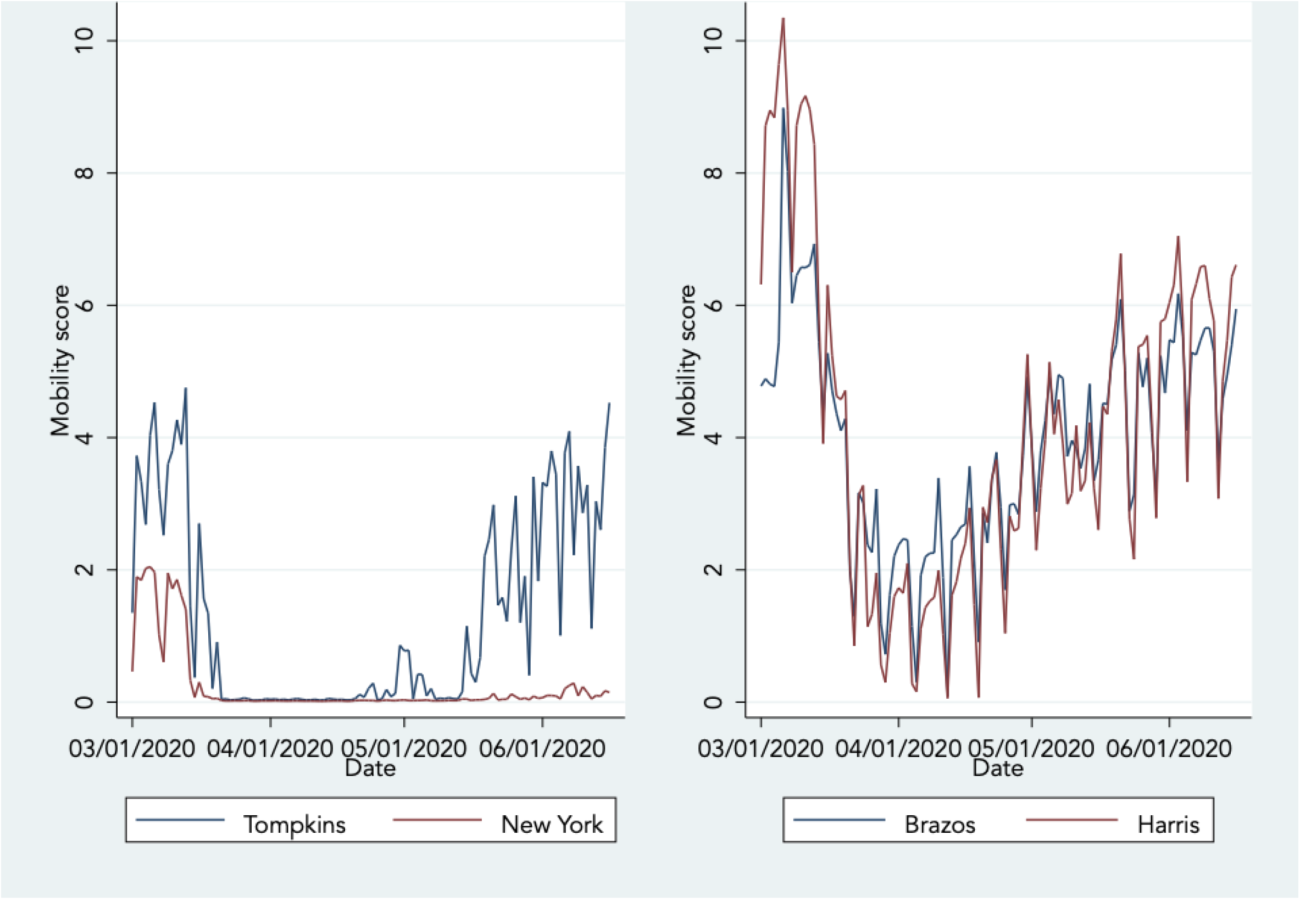
Social Mobility Index. This figure shows the evolution of the social mobility index from March 1, 2020, to the end of our sample period for Tompkins and New York counties in the state of New York, and for Brazos and Harris counties in the states of Texas. We selected these counties and states at random for illustrative purposes.

Finally, we construct a comprehensive list of protests that took place across the U.S. Our starting point is the List of George Floyd protests in the United States assembled by Wikipedia. At the time of writing, the main Wikipedia page cited 134 news articles from national, regional, and local media outlets, and the secondary pages cited hundreds more. From these media citations, we extracted the location and the date at which the protests reportedly took place, as well as the estimated number of individuals involved in each protest. We complement this process with a search on the Dow Jones Factiva database.

### 2.2 Regression Specification

We examine the impact of the abrupt relaxation of social distancing practices, which occurred during the U.S. nationwide protests, on the SARS-CoV-2 infection rate with an Ordinary Least Squares (OLS) differences-in-differences (DID) panel regression equation, which is specified as follows:

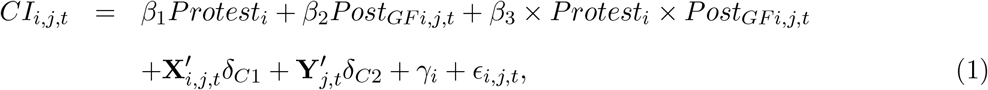

where *CI*_*i,j,t*_ corresponds to new confirmed SARS-CoV-2 infections in county *i* from state *j* on day *t*, per 100,000 population. *Protest*_*i*_ is an indicator variable which is set equal to one if a protest took place in county *i*, and to zero otherwise. *Post*_*GF i,j,t*_ is an indicator variable set equal to zero from the first day of our sample period up until May 25, 2020, the day of George Floyd’s tragic death, and to one on every subsequent date. *Protest × Post*_*GF*_ is an indicator variable which captures the interaction between *Protest*_*i*_ and *Post*_*GF i,j,t*_. **X**_*i,j,t*_ and **Y**_*j,t*_ are vectors of county and state characteristics which we use as control variables, and (*γ*_*i*_) represents state-level fixed effects to control for time-invariant differences across states in our regressions.

In equation (1), *β*_1_ captures any differences that may exist between the SARS-CoV-2 infection rate in protest and non-protest counties, and that are unrelated to the protests. We expect this coefficient to be statistically indistinguishable from zero. *β*_2_ captures the impact of the relaxation of social distancing practices on the infection rate across all U.S. counties, following the onset of the protests. *β*_3_ captures the incremental impact of the relaxation of social distancing practices on the infection rate, specifically in counties where protests took place. Under the null hypothesis that the relaxation of social distancing practices has no causal impact on the SARS-CoV-2 infection rate, both *β*_2_ and *β*_3_ coefficients should be statistically indistinguishable from zero. In all our regressions, we cluster the standard errors at the county level to account for any potential cross-sectional dependence in the error terms, *E*_*i,j,t*_.^17^ We perform our statistical analysis with STATA 16 and use Sergio Correia’s REGHDFE command to estimate equation (1).^18^

### 2.3 Research Setting

Two key requirements for the identification of the causal link between social distancing and the spread of SARS-CoV-2 are satisfied in our research setting, namely: 1) the existence of a strong theoretical basis supporting the relationship in question and, 2) exogenous variation in the variable of interest, i.e social distancing. ^19^ The latter is key to establish causality, because it mitigates concerns that omitted variables correlated with both the protests and the spread of SARS-CoV-2 might be driving our findings. This experimental setting also enables us to circumvent common concerns about endogeneity and self-selection which besets most non-randomized-trial experiments.^20^

### 2.4 Covariates

In our differences-in-differences regressions, we include control variables which may influence the transmission rate of SARS-CoV-2. These control variables account for demographic, health, geographic, and income level variations across counties. For demographic indicators, we include male sex and age (60 years+) since these factors are associated with both an increased risk of testing positive for SARS-CoV-2 and greater illness severity.^21^ We also include ethnicity as a demographic variable to account for the increased risk of a positive SARS-COV-2 test observed among Blacks and Hispanics. Obesity, diabetes, and hypertension are clinical risk factors included as health covariates in the regressions, as they are associated with an increased risk of severe illness, and a greater risk of mortality from COVID-19. ^22^ We also include smoking as a clinical risk factor, as some evidence suggests that smoking may be associated with an increased severity of COVID-19. ^23^ We include population density among our control variables, as higher rates of SARS-CoV-2 infections are observed in more densely populated, urban areas. ^22,24^ Consistent with previous research showing that residents from more economically deprived areas are more likely to test positive for SARS-COV-2, we use real GDP per capita to control for income in our regressions. ^22^

## 3 Results

### 3.1 Impact of Protests on SARS-CoV-2 Infections

We report results from regression equation (1) in Table III. In Model (1), the coefficients associated with *Protest* is equal to 1·22 (95%CI: 0·79–1·65) and is highly significant. This implies that, over the entire sample period, the SARS-CoV-2 infection rate is 1·22 cases per day, per 100,000 population higher in the counties where protests took place, relative to the counties where no protests took place. The coefficient associated with *Post*_*GF*_ is positive and highly significant, implying that the SARS-CoV-2 infection rate increases by 3·39 cases per day, per 100,000 across the U.S. following the onset of the protests. Finally, the coefficient associated with the *Protest × Post*_*GF*_ interaction indicates that the infection rate is even greater in the counties in which protests actually took place, following the onset of the protests (4·01; 95%CI: 3·24–4·78). To put this number into perspective, recall that the average number of new cases across all counties is equal to 5 per day, per 100,000 population, in the week preceding the onset of the protests (see Column (2) of Table I). Using this number as a reference point, COVID-19 cases increase by a further 80·2%, on average, in protest-counties, relative to non-protest counties, and by 3·39 + 4·01 = 7·40 cases per day, per 100,000 population, or 148% overall. Models (2)-(6) of Table III provide evidence that is consistent with Model (1). The coefficient associated with *Protest* loses its statistical significance in Models (2) and (6), suggesting that the higher overall infection rate of protest-counties is attributable to cross-county differences in demography. We note that the coefficient associated with *Post*_*GF*_ is very stable across the six models, ranging between 3·33 and 3·39. Likewise, the coefficient associated with our *Protest × Post*_*GF*_ interaction is quite stable across the six models, ranging between 2·80 in Model (6) and 4·01 in Model (1). Although there is no telling which one of these six models provides a better description of the causal impact of relaxing social distancing practices on the spread of SARS-CoV-2, out of conservatism, we will employ our omnibus regression Model (6) from this point on, referring to this specification as our baseline regression model.

**Table II:** List of U.S. Protests. This table reports the earliest and the latest date at which the first protest took place in any county within a particular state, as well as the smallest and the largest number of participants reported to have taken part in this first protest.

| State | First date |  | Number of participants |  |
| --- | --- | --- | --- | --- |
|  | Earliest | Latest | Smallest | Largest |
| Alabama | 2020-05-29 | 2020-06-01 | 50 | 1,000 |
| Alaska | 2020-05-30 | 2020-06-06 | 20 | 1,400 |
| Arizona | 2020-01-06 | 2020-05-30 | 50 | 1,000 |
| Arkansas | 2020-05-30 | 2020-06-01 | 100 | 1,000 |
| California | 2020-05-28 | 2020-06-03 | 100 | 3,000 |
| Colorado | 2020-05-28 | 2020-06-04 | 50 | 1,000 |
| Connecticut | 2020-05-29 | 2020-05-31 | 100 | 1,000 |
| Delaware | 2020-05-30 | 2020-06-01 | 30 | 1,000 |
| District of Columbia | 2020-05-29 | 2020-05-29 | 1,000 | 1,000 |
| Florida | 2020-05-29 | 2020-06-06 | 30 | 1,200 |
| Georgia | 2020-05-29 | 2020-06-01 | 50 | 1,000 |
| Hawaii | 2020-05-30 | 2020-05-30 | 100 | 150 |
| Idaho | 2020-05-30 | 2020-06-03 | 25 | 1,000 |
| Illinois | 2020-05-29 | 2020-06-06 | 15 | 1,400 |
| Indiana | 2020-05-29 | 2020-06-04 | 100 | 10,000 |
| Iowa | 2020-05-29 | 2020-06-05 | 20 | 1,000 |
| Kansas | 2020-05-30 | 2020-06-06 | 25 | 2,000 |
| Kentucky | 2020-05-28 | 2020-05-31 | 100 | 1,000 |
| Louisiana | 2020-05-29 | 2020-06-04 | 25 | 1,000 |
| Maine | 2020-05-29 | 2020-06-07 | 100 | 1,000 |
| Maryland | 2020-05-29 | 2020-06-03 | 100 | 1,000 |
| Massachusetts | 2020-05-28 | 2020-06-02 | 25 | 5,000 |
| Michigan | 2020-05-28 | 2020-06-01 | 100 | 5,000 |
| Minnesota | 2020-05-26 | 2020-06-02 | 100 | 5,000 |
| Mississippi | 2020-05-28 | 2020-05-30 | 25 | 1,000 |
| Missouri | 2020-05-29 | 2020-06-07 | 100 | 2,000 |
| Montana | 2020-05-29 | 2020-05-31 | 50 | 1,000 |
| Nebraska | 2020-05-29 | 2020-06-03 | 20 | 5,000 |
| Nevada | 2020-05-29 | 2020-06-06 | 20 | 1,000 |
| New Hampshire | 2020-05-30 | 2020-06-03 | 100 | 1,000 |
| New Jersey | 2020-05-30 | 2020-06-06 | 35 | 10,000 |
| New Mexico | 2020-05-28 | 2020-06-01 | 40 | 1,000 |
| New York | 2020-05-28 | 2020-06-07 | 100 | 11,000 |
| North Carolina | 2020-05-29 | 2020-06-04 | 25 | 1,000 |
| North Dakota | 2020-05-30 | 2020-06-04 | 50 | 1,000 |
| Ohio | 2020-05-28 | 2020-06-05 | 30 | 5,000 |
| Oklahoma | 2020-05-30 | 2020-06-03 | 1,000 | 1,000 |
| Oregon | 2020-05-28 | 2020-06-04 | 10 | 2,000 |
| Pennsylvania | 2020-05-30 | 2020-06-07 | 15 | 5,000 |
| Rhode Island | 2020-05-30 | 2020-06-06 | 100 | 1,000 |
| South Carolina | 2020-05-30 | 2020-05-31 | 300 | 1,000 |
| South Dakota | 2020-05-29 | 2020-06-05 | 30 | 1,000 |
| Tennessee | 2020-05-27 | 2020-05-31 | 50 | 5,000 |
| Texas | 2020-05-29 | 2020-06-06 | 50 | 5,000 |
| Utah | 2020-05-30 | 2020-05-31 | 100 | 1,000 |
| Vermont | 2020-05-30 | 2020-06-03 | 100 | 1,200 |
| Virginia | 2020-05-29 | 2020-06-07 | 15 | 1,500 |
| Washington | 2020-05-29 | 2020-06-05 | 100 | 2,000 |
| West Virginia | 2020-05-30 | 2020-05-31 | 50 | 1,000 |
| Wisconsin | 2020-05-29 | 2020-06-03 | 100 | 1,000 |
| Wyoming | 2020-05-29 | 2020-06-03 | 10 | 1,000 |

**Table III:**
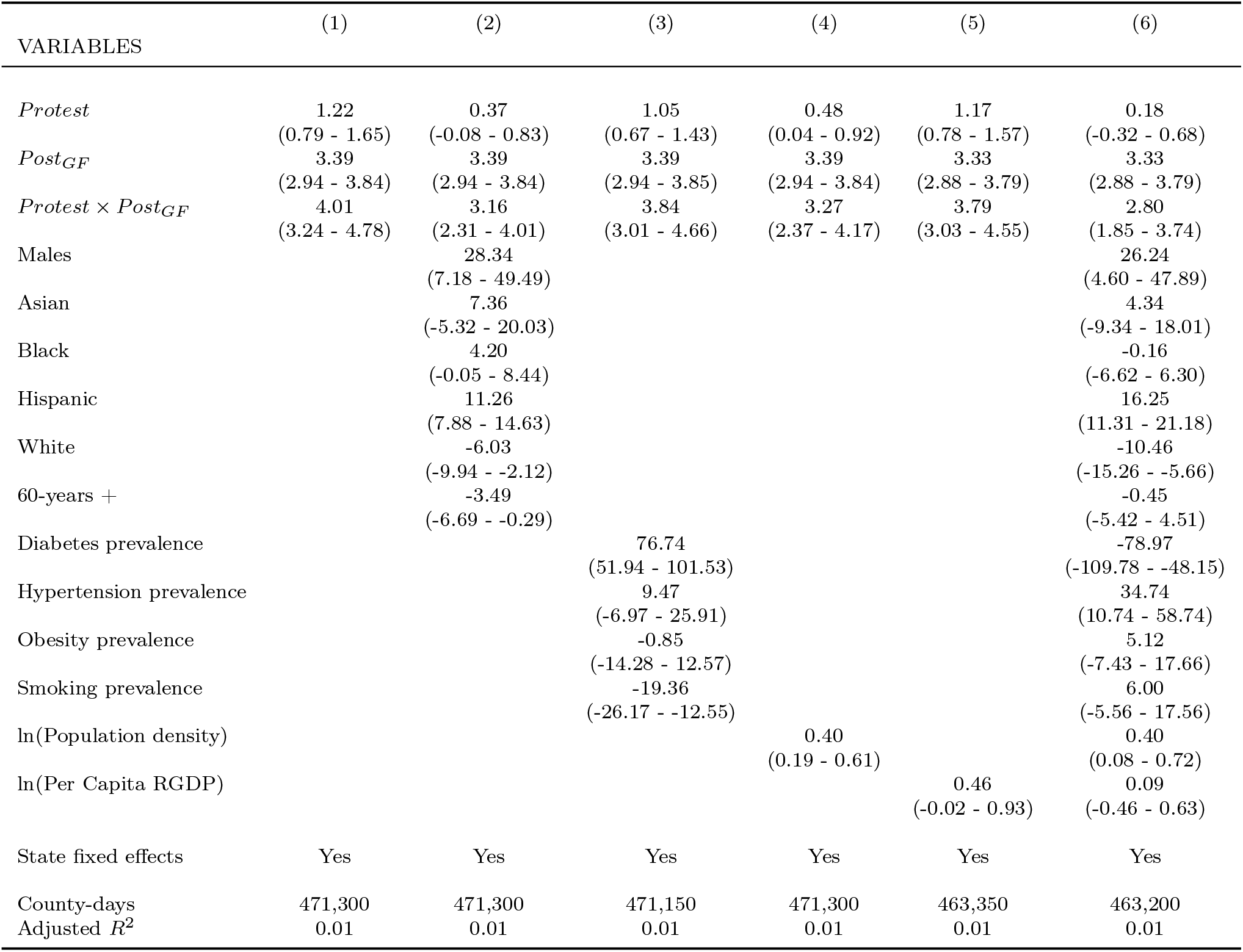
Impact of Protests on SARS-CoV-2 Infections. This table reports results from our differences-in-differences regression equation (1). In these regressions, the dependent variable corresponds to the county-level number of new confirmed COVID-19 cases, per day, per 100,000 population. *Protest* is an indicator variable set equal to one in counties where protests reportedly took place, and to zero in counties where no protests took place. *Post*_*GF*_ is an indicator variable set equal to zero up until George Floyd’s tragic death, and to one on every subsequent date. The 95% confidence intervals reported under the regression coefficients are based on standard errors that are clustered at the county level. ^17^

Based on the evidence reported in Table III, we can safely reject the null hypothesis that relaxing social distancing practices has no impact on the spread of SARS-CoV-2. Furthermore, as our placebo tests (Sub-section 3.3) indicate, we can rule out the possibility that this important finding is attributable to chance with a high degree of confidence.

### 3.2 Social Distancing Restriction and Mobility

In the period preceding the onset of the protests, the number of new COVID-19 cases began to drop steadily across the country.^3^ Accordingly, several states began to unwind their social distancing restrictions in a carefully staged manner. Figure (2) illustrates this trend in Alabama, California, Florida, and New York, for instance. Starting in mid-March, we observe a steady rise in our social distancing restrictions index in these four states and we observe the start of a slow unwind by mid-April. Notably, while social distancing restrictions were being relaxed across the nation, social mobility was on the rise (see Figure 3). Consequently, it may very well be that the concurrent relaxation of social distancing restrictions and the increase in social mobility during the event period has prompted individuals to relax their social distancing practices, and that the effect that we document in Table III is partly contaminated by these contemporaneous changes. We address this issue in Table IV, where we include our social distancing restrictions and social mobility indexes in our baseline DID regression equation (1) as additional control variables.

**Table IV:** Social Distancing Restrictions and Social Mobility. This table evaluates the impact of the concurrent relaxation of state-imposed social distancing restrictions and social mobility on the causal link between social distancing practices and SARS-CoV-2 infection rate documented in Table III. Model (1) corresponds to our baseline regression Model (6) from Table III. Model (2) includes our social distancing restrictions index as a supplementary control variable, Model (3) includes our social mobility index as a supplementary control variable, and Model (4) includes both supplementary variables. The 95% confidence intervals reported under the regression coefficients are based on standard errors that are clustered at the county level. ^17^

| VARIABLES | (1) | (2) | (3) | (4) |
| --- | --- | --- | --- | --- |
| *Protest* | 0.18<br>(-0.32 - 0.68) | 0.22<br>(-0.28 - 0.72) | 0.29<br>(-0.49 - 1.06) | 0.37<br>(-0.40 - 1.14) |
| *Post<sub>GF</sub>* | 3.33<br>(2.88 - 3.79) | 3.01<br>(2.55 - 3.47) | 2.97<br>(2.38 - 3.56) | 3.06<br>(2.47 - 3.65) |
| *Protest* × *Post<sub>GF</sub>* | 2.80<br>(1.85 - 3.74) | 2.29<br>(1.33 - 3.26) | 1.81<br>(0.67 - 2.95) | 1.73<br>(0.59 - 2.87) |
| Social distancing restrictions |  | 0.51<br>(0.47 - 0.56) |  | 0.50<br>(0.44 - 0.55) |
| Social mobility |  |  | -0.73<br>(-0.91 - -0.55) | -0.26<br>(-0.45 - -0.08) |
| Males | 26.24<br>(4.60 - 47.89) | 26.24<br>(4.60 - 47.89) | 48.62<br>(0.66 - 96.58) | 48.07<br>(0.09 - 96.05) |
| Asian | 4.34<br>(-9.34 - 18.01) | 4.34<br>(-9.34 - 18.01) | 8.41<br>(-17.58 - 34.41) | 10.77<br>(-15.24 - 36.79) |
| Black | -0.16<br>(-6.62 - 6.30) | -0.16<br>(-6.62 - 6.30) | -7.49<br>(-22.19 - 7.20) | -7.31<br>(-22.04 - 7.42) |
| Hispanic | 16.25<br>(11.31 - 21.18) | 16.25<br>(11.31 - 21.18) | 24.67<br>(16.99 - 32.35) | 25.24<br>(17.55 - 32.93) |
| White | -10.46<br>(-15.26 - -5.66) | -10.46<br>(-15.26 - -5.66) | -21.75<br>(-34.01 - -9.48) | -22.19<br>(-34.52 - -9.87) |
| 60-years + | -0.45<br>(-5.42 - 4.51) | -0.45<br>(-5.42 - 4.51) | 0.28<br>(-7.26 - 7.81) | 1.13<br>(-6.45 - 8.70) |
| Diabetes prevalence | -78.97<br>(-109.78 - -48.15) | -78.97<br>(-109.78 - -48.15) | -126.67<br>(-176.41 - -76.93) | -124.31<br>(-173.93 - -74.68) |
| Hypertension prevalence | 34.74<br>(10.74 - 58.74) | 34.74<br>(10.74 - 58.74) | 58.01<br>(17.42 - 98.59) | 55.94<br>(15.36 - 96.52) |
| Obesity prevalence | 5.12<br>(-7.43 - 17.66) | 5.12<br>(-7.43 - 17.66) | 5.10<br>(-14.50 - 24.69) | 3.35<br>(-16.27 - 22.96) |
| Smoking prevalence | 6.00<br>(-5.56 - 17.56) | 6.00<br>(-5.56 - 17.56) | 12.66<br>(-6.28 - 31.61) | 13.73<br>(-5.22 - 32.68) |
| ln(Population density) | 0.40<br>(0.08 - 0.72) | 0.40<br>(0.08 - 0.72) | 0.36<br>(-0.19 - 0.92) | 0.43<br>(-0.13 - 0.98) |
| ln(Per Capita RGDP) | 0.09<br>(-0.46 - 0.63) | 0.09<br>(-0.46 - 0.63) | -0.21<br>(-1.11 - 0.68) | -0.19<br>(-1.08 - 0.71) |
| State fixed effects | Yes | Yes | Yes | Yes |
| County-days | 463,200 | 463,200 | 279,754 | 279,754 |
| Adjusted $R^2$ | 0.01 | 0.02 | 0.02 | 0.02 |

Table IV includes four models. Model (1) corresponds to Model (6) from Table III, Model (2) includes the additional control for social distancing restrictions, Model (3) includes the additional control for social mobility, and Model (4) includes both controls. In Model (2), we see a drop from 3·33 to 3·01 in the coefficient associated with *Post*_*GF*_, relative to Model (1), but the coefficient remains highly significant. We observe a similar drop in the coefficient associated with the *Protest× Post*_*GF*_ interaction, from 2·80 to 2·29, with no drop in its statistical significance.

In Model (3), controlling for social mobility has a slightly larger impact on the coefficients associated with *Post*_*GF*_ and *Protest × Post*_*GF*_. The first coefficient drops from 3·33 to 2·97, while the *Protest × Post*_*GF*_ drops from 2·80 to 1·81. Evidently, social distancing restrictions and social mobility are correlated with one another. For instance, we should expect social mobility to rise when travel restrictions are lifted. When we control for both factors in Model (4), the coefficient associated with *Post*_*GF*_ is equal to 3·06 (95%CI: 2·47–3·65), which is highly significant, and the coefficient associated with *Protest×Post*_*GF*_ is equal to 1·73 (95%CI: 0·59–2·87), also highly significant.

In summary, after controlling for the reduction in social distancing restrictions and the increase in social mobility that occurred following the onset of the protests, we still observe a significant increase in the number of daily COVID-19 cases across all counties (61·2% relative to the week preceding the event), and a further increase of 1·73 cases (34·6%) in the counties where protests took place. We attribute the latter to the relaxation of social distancing practices during the protests. This interpretation is supported by the abundance of video footage demonstrating that the mass protests brought people into close physical proximity to one another, in contravention to social distancing restrictions that were in place at the time.

### 3.3 Placebo Test

In Table V, we report the results of a placebo test assessing whether the causal impact of the protests on the spread of SARS-CoV-2 that we document in Tables III and IV can be attributed to chance. For this purpose, we implement a Monte Carlo simulation exercise centered on our baseline DID panel regression specification (1), i.e. Model (4) of Table IV. In this simulation, we pick a random date between February 6, 2020, and June 1, 2020, to represent the onset of the protests and we assign counties to the protest group randomly, in proportion to the actual fraction of counties that took part in the protests (18%). We carry out this exercise 10,000 times, and each time, we estimate our model with the simulated protest onset date and protest county pair, and collect the key parameter estimates from the regression, i.e. *Protest, Post*_*GF*_, and *Protest × Post*_*GF*_, along with their county-cluster robust *t*-statistics.

**Table V:**
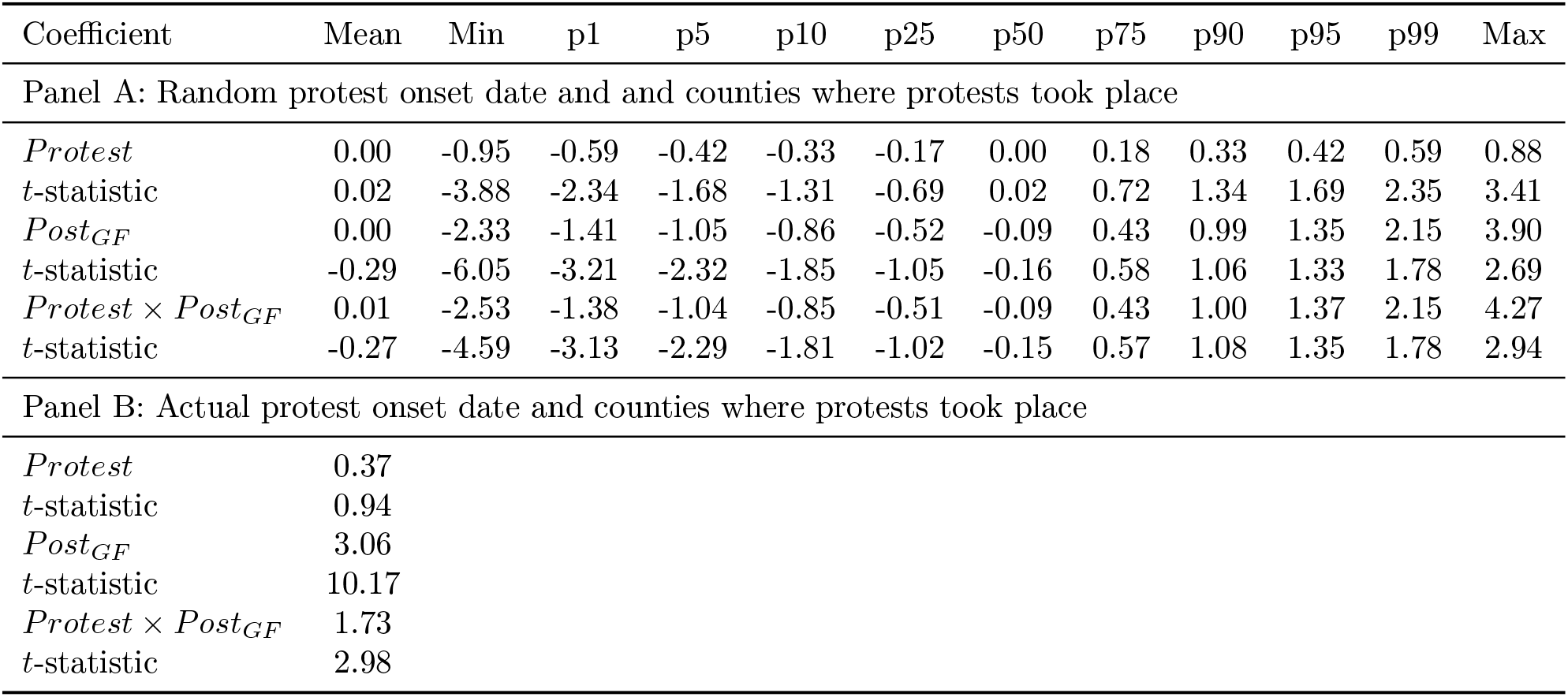
Placebo Tests. This table reports results from a Monte Carlo simulation of the impact of the protests on the SARS-CoV-2 infection rate across the U.S. This analysis is centered on our baseline differences-in-differences panel regression specification, i.e. Model (4), presented in Table IV. In this simulation, we estimate our baseline regression model 10,000 times. In each regression, we assign a random date for the start of the protests, ranging between February 6, 2020, and June 1, 2020, and we assign the counties to the group of protest participants randomly, in proportion to the actual fraction of counties that took part in the protests (18%). In Panel A, we report the simulated distribution of the regression model’s key parameter estimates, i.e. *Protest, Post*_*GF*_, and *Protest* x *Post*_*GF*_, along with the distribution of the t-statistics for these coefficients. In Panel B, we report the actual value of the parameter estimates from Model (6) of Table III to facilitate comparisons. The *t*-statistics are based on standard errors that are robust to country-level clusters. ^17^

In this simulation, the impact of the random date on the randomly assigned protest counties on the SARS-CoV-2 infection rate is negligible, on average. Only 25% of the coefficients from the simulation are positive, and at most 1% of them are statistically significant. Furthermore, the coefficients associated with *Post*_*GF*_ and *Protest × Post*_*GF*_ from the actual regressions (Panel B), i.e. 3·06 and 1·73, are well above the 95% confidence thresholds inferred from the simulated distribution, i.e. 1·35 and 1·37, respectively. Indeed, our baseline regression coefficients fall at the very top end of the simulated distribution. This implies that we can safely reject the null hypothesis that the causal impact of protests on the SARS-CoV-2 infection rate that we document is due to pure chance, with at least 99% confidence.

## 4 Discussion

In this paper, we employ the natural experimental setting created by the U.S. protests precipitated by George Floyd’s tragic death to document the causal impact of social distancing measures on the spread of SARS-CoV-2. Using a DID analysis, in which the treatment effect corresponds to the onset of the protests and the treatment group corresponds to the counties in which protests reportedly took place, we document a country-wide increase of more than 3·06 cases per day, per 100,000 population, following the onset of the protests, and a further increase of 1·73 cases per day, per 100,000 population, in the counties in which the protests took place. Relative to the average number of new COVID-19 cases per day during the week preceding the onset of the protests, this represents a 61·2% country-wide increase in COVID-19 cases, and further 34·6% increase in the protest counties.

### Strengths and Weaknesses

Early predictive models assessing the effectiveness of social distancing have suggested that a greater spread of SARS-CoV-2 would occur in the absence of social distancing measures.^25–27^ Similarly, our study demonstrates that when social distancing is reduced, i.e by individuals protesting in close proximity, the spread of SARS-CoV-2 increases. Our study differs from its predecessors because instead of examining the effectiveness of social distancing measures following their imposition, ^11,12,14^ we examine the impact of social distancing on the spread of COVID-19 when social distancing practices are abruptly relaxed. Additionally, unlike previous studies, we do not use mobility as a measure of social distancing, instead we control for social mobility as a variable in our analyses. By explicitly controlling for the concurrent increase in social mobility and the relaxation of state-imposed social distancing restrictions during the period surrounding the protests, our study demonstrates that social distancing directly impacts the spread of SARS-CoV-2. We also control for a host of covariates known to influence the transmission of SARS-CoV-2, and implement placebo tests to rule out the possibility that our results are attributable to chance. Therefore, we can be confident that the increase in SARS-CoV-2 infections that we observe following the onset of the protests can be attributed to the relaxation of social distancing practices.

Our study is not without limitations. In particular, over 70 testing centers across the U.S. were closed following the onset of the protests. Therefore, the increase in the SARS-CoV-2 infections that we document likely underestimates the true increase. We are also unable to assess protest participants’ vulnerability (e.g. age, underlying health conditions, personal protective wear, etc.), and variability along these dimensions may influence the risk of SARS-CoV-2 infection. Additionally, we cannot control for the actual degree of physical proximity between participants, which would impact the transmission rate of SARS-CoV-2 during the protests. Moreover, we rely on the accuracy of media reports to identify the counties in which protests took place. Finally, we do not account for the magnitude of the protests in each county, however, expressing the case counts in rates rather than in levels should minimize any potential scale-related effects.

### Future Research and Implications

Future research using this experimental setting could use machine learning tools to analyze protest videos and determine the relative contribution of participant demographics, the degree of physical distancing, and the extent and type of personal protective wear on the spread of SARS-CoV-2. Taken together, this study demonstrates that, when controlling for social mobility and restrictions, social distancing practices causally impact the spread of SARS-CoV-2. As states are in the midst of relaxing the social distancing restrictions initially imposed in March 2020, establishing the effectiveness of social distancing practices in a statistically reliable way has important public health implications. Our research informs policy makers and provides insights regarding the usefulness of social distancing as an intervention to minimize the spread of SARS-CoV-2, and reduce the risk of a second, and possibly, third wave of COVID-19.

## Data Availability

This study uses publicly accessible data exclusively.

https://github.com/descarteslabs/DL-COVID-19

https://github.com/COVID19StatePolicy/SocialDistancing

http://ghdx.healthdata.org/record/ihme-data/united-states-hypertension-estimates-county-2001-2009

http://ghdx.healthdata.org/record/ihme-data/united-states-smoking-prevalence-county-1996-2012

http://ghdx.healthdata.org/record/ihme-data/united-states-diabetes-prevalence-county-1999-2012

http://ghdx.healthdata.org/record/ihme-data/united-states-physical-activity-and-obesity-prevalence-county-2001-2011

https://github.com/CSSEGISandData/COVID-19/tree/master/csse_covid_19_data/csse_covid_19_time_series

https://apps.bea.gov/regional/downloadzip.cfm

https://www2.census.gov/programs-surveys/popest/datasets/2010-2019/counties/totals/co-est2019-alldata.csv

## Acknowledgments

We wish to express our sincere thanks to the Descartes Labs for making their mobility data available to us.

## Contributors

LG conceived the study, and all authors contributed to the final study design. LG performed the data analysis, created the tables and figures, and wrote the methods and results sections in the initial draft of the manuscript. SG and JL conducted the literature search, and assisted LG with the data collection. All authors contributed substantially to the interpretation of the data, and equally to the write up. All authors wrote and approved of the final manuscript submission. The corresponding author attests that all listed authors meet authorship criteria and that no others meeting the criteria have been omitted.

## Ethical Approval

Not required. This study uses existing data sources that are fully publicly available and anonymized.

## Data Sharing

This study uses publicly accessible data exclusively.

**Summary Box**

### What is already known on this topic

- Prior studies suggest that there as an association between social distancing and the spread of SARS-CoV-2 following the imposition of social distancing measures
- The effectiveness of social distancing is generally examined using mobility as a proxy for social distancing

### What this study adds

- The causal link between social distancing practices and the spread of SARS-Cov-2 is established by exploiting a purely exogenous shock, i.e the protests, in a framework that explicitly controls for social mobility and social distancing restrictions
- Placebo tests rule out the possibility that the causal link documented herein is due to chance

